# A Tool to Early Predict Severe Corona Virus Disease 2019 (COVID-19) : A Multicenter Study using the Risk Nomogram in Wuhan and Guangdong, China

**DOI:** 10.1101/2020.03.17.20037515

**Authors:** Jiao Gong, Jingyi Ou, Xueping Qiu, Yusheng Jie, Yaqiong Chen, Lianxiong Yuan, Jing Cao, Mingkai Tan, Wenxiong Xu, Fang Zheng, Yaling Shi, Bo Hu

## Abstract

**Background:** Due to no reliable risk stratification tool for severe corona virus disease 2019 (COVID-19) patients at admission, we aimed to construct an effective model for early identifying cases at high risk of progression to severe COVID-19.

**Methods:** In this retrospective three-centers study, 372 non-severe COVID-19 patients during hospitalization were followed for more than 15 days after admission. Patients who deteriorated to severe or critical COVID-19 and patients who kept non-severe state were assigned to the severe and non-severe group, respectively. Based on baseline data of the two groups, we constructed a risk prediction nomogram for severe COVID-19 and evaluate its performance.

**Results:** The train cohort consisted of 189 patients, while the two independent validation cohorts consisted of 165 and 18 patients. Among all cases, 72 (19.35%) patients developed severe COVID-19. We found that old age, and higher serum lactate dehydrogenase, C-reactive protein, the coefficient of variation of red blood cell distribution width, blood urea nitrogen, direct bilirubin, lower albumin, are associated with severe COVID-19. We generated the nomogram for early identifying severe COVID-19 in the train cohort (AUC 0.912 [95% CI 0.846-0.978], sensitivity 85.71%, specificity 87.58%); in validation cohort (0.853 [0.790-0.916], 77.5%, 78.4%). The calibration curve for probability of severe COVID-19 showed optimal agreement between prediction by nomogram and actual observation. Decision curve and clinical impact curve analysis indicated that nomogram conferred high clinical net benefit.

**Conclusion:** Our nomogram could help clinicians to early identify patients who will exacerbate to severe COVID-19, which will enable better centralized management and early treatment of severe patients.

**Summary:** Older age; higher LDH, CRP, RDW, DBIL, BUN; lower ALB on admission correlated with higher odds of severe COVID-19. An effective prognostic nomogram composed of 7 features could allow early identification of patients at risk of exacerbation to severe COVID-19.

## Introduction

Since the outbreak of novel coronavirus pneumonia (COVID-19) in December 2019, the number of reported cases has surpassed 260,000 with over 11180 deaths worldwide, as of March 22 2020. The severe acute respiratory syndrome coronavirus 2 (SARS-CoV-2), a member of coronaviruses known to cause common colds and severe illnesses such as, is the cause of COVID-19[1]. Compared with much higher overall case-fatality rates (CFR) for the severe acute respiratory syndrome (SARS) and middle east respiratory syndrome (MERS), COVID-19 is being responsible for more total deaths because of the increased transmission speed and the growing numbers of cases [2]. Up to now, the World Health Organization (WHO) has raised global Coronavirus Disease (COVID-19) outbreak risk to “Very High”, and SARS-CoV-2 infection has become a serious threat to public health.

According to a report recently released by the Chinese Center for Disease Control and Prevention (CDC) that included approximately 44,500 confirmed cases of SARS-CoV-2 infections, up to 15.8% were severe or critical. Most COVID-19 patients have a mild disease course, while some patients experience rapid deterioration (particularly within 7-14 days) from onset of symptoms into severe COVID-19 with or without acute respiratory distress syndrome (ARDS)[3]. Current epidemiological data suggests that the mortality rate of severe COVID-19 patients is about 20 times higher than that of non-severe COVID-19 patients [4]. This situation highlights the need to identify COVID-19 patients at risk of approaching to severe COVID-19. These severe illness patients often require utilization of intensive medical resources. Therefore, early identification of patients at high risk for progression to severe COVID-19 will facilitate appropriate supportive care and reduce the mortality rate, unnecessary or inappropriate healthcare utilization via patient prioritization.

At present, an early warning model for predicting COVID-19 patients at-risk of developing a costly condition is scarce [3, 5]. So far, prognosis factors of COVID-19 mainly focus on the immune cells. In our study, we found that older age, higher lactate dehydrogenase (LDH) and C-reactive protein (CRP), RDW (the coefficient of variation of red blood cell distribution width), DBIL (direct bilirubin), blood urea nitrogen (BUN), and lower albumin (ALB) on admission correlated with higher odds of severe COVID-19. Based on these indexes, we developed and validated an effective prognostic nomogram with high sensitivity and specificity for accurate individualized assessment of the incidence of severe COVID-19. Among these indexes, the prognostic role of RDW in COVID-19 is underestimated, which is associated with the increased turnover of erythrocytes. Our results hinted that the turnover of RBC might involve in severe illness.

## Material and method

### Data collection

Data on COVID-19 inpatients between January 20^th^ 2020 and March 2^nd^ 2020 was retrospectively collected from three clincial centers: Guangzhou Eighth People’s Hospital, Zhongnan Hospital of Wuhan University and the Third Affiliated Hospital of Sun Yat-sen University. A total of 381 patients with COVID-19 were enrolled, 9 patients younger than 15 years of age were excluded from the study. Clinical laboratory test results, including SARS-CoV-2 RNA detection results, biochemical indices, blood routine results, were collected from routine clinical practice. Written informed consent was waived by the Ethics Commission of each hospital for emerging infectious diseases. The study was approved by the Ethics Committee of the Eighth People’s Hospital of Guangzhou, the Ethics Commission of the Third Affiliated Hospital of Sun Yat-sen University and the Ethics Commission of Zhongnan Hospital.

The diagnosis of SARS-CoV-2 infection was based on the Guidelines for Diagnosis and Treatment of Novel Coronavirus Pneumonia (5^th^ version), released by National Health Commission of China. Suspected cases of COVID-19 requires meeting any of the following epidemiology history criteria or any two of the following clinical manifestations: (A) Epidemiological history: a history of travel to or residence in Wuhan in the last 14 days prior to symptom onset; contact with a confirmed or suspected case of 2019-nCOV infection in the last 14 days prior to symptom onset; aggressive disease onset. (B) Clinical manifestation: Fever and/or respiratory infection, or with normal/decreased white blood cells counts and normal/decreased lymphocyte counts. In the absence of the above mentioned criteria for epidemiological history, the suspected case should meet with all of the above mentioned criteria for clinical manifestation. A confirmed case was defined as an individual with laboratory confirmation of SARS-CoV-2 which required positive results of SARS-CoV-2 RNA, irrespective of clinical signs and symptoms. For diagnosis of Severe COVID-19 group, at least one of the following conditions should be met: (1) Shortness of breath, Respiratory rate (RR) ≥ 30times/min, (2) Arterial oxygen saturation (Resting status) ≤93%, or (3) the ratio of Partial pressure of oxygen to Fraction of inspiration O-2(PaO-2/ FiO-2)≤300mmHg.

### Laboratory Methods

Clinical laboratory test results, including SARS-CoV-2 RNA detection results, biochemical indices, blood routine results, were collected from routine clinical practice. Clinical laboratory test results included albumin (ALB), aspartate aminotransferase (AST), alanine transaminase (ALT), blood urea nitrogen (BUN), creatine kinase (CK), creatine kinase-MB (CK-MB), creatinine (Crea), C-reactive protein (CRP), total bilirubin (TBIL), direct bilirubin (DBIL), globulin (GLB), lactate dehydrogenase (LDH), procalcitonin (PCT), total bile acid (TBA), hemoglobin (HB), lymphocyte count, monocyte count, neutrophil count, platelet distribution width (PDW), platelet (PLT), red blood cell (RBC), RDW (red blood cell distribution width-coefficient variation). SARS-CoV-2 RNA were detected using real time quantitive PCR (RT-qPCR) on nucleic acid extracted from upper respiratory swab samples. Upper respiratory swab samples were collected on all suspected cases of SARS-CoV-2 infection on admission and immediately placed into sterile tubes with viral transport medium. All biochemical and hematology parameters were obtained via standard automated laboratory methods and using commercially available kits following to the manufacturers protocols.

### Statistical Analysis

Categorical variables were expressed as frequency and percentages, and Fisher’s exact test was performed to analyze the significance. Continuous variables were expressed as mean (standard deviation [SD]), or median (interquartile range [IQR]), as appropriate. Parametric test (T test) and non-parametric test (Mann-Whitney U) were used for continuous variables with or without normal distribution, respectively. A value of p < 0.05 was considered statistically significant. Except for filling missing values, all the statistical analyses were analyzed using R (version 3.6.2) with default parameters.

Of all potential predictors in the dataset, 0.09 % of the fields had missing values. Predictor exclusion was limited to those with more than 7% missing rate to minimize the bias of the regression coefficient. [16]. Little’s MCAR test (R package BaylorEdPsych) was used to assess the suitability of the remaining missing values for imputation. This test is used to test whether missing values are “missing completely at random” (MCAR) or biased. The missing values were imputed by expectation-maximization (EM) method using SPSS statistical software, version 25 (SPSS, Inc., Chicago, IL, USA).

To identify the relative importance of each feature, feature selection was performed using the least absolute shrinkage and selection operator (LASSO) regression method, and prediction models were built using logistic regression, decision tree, random forest (RF) and support vector machine (SVM) using R package mlr, using 3-fold cross-validation for diverse parameter conditions, respectively. As described previously, Nomograms were established with the rms package and the performance of nomogram was evaluated by area under the receiver operating characteristic curve (AUC) and calibration (calibration plots and Hosmer-Lemeshow calibration test) in R. During the external validation of the nomogram, the total points for each patient in the validation cohort were calculated based on the established nomogram.

## Results

### Clinicopathologic characteristics of COVID-19 patients

The selection of the study population is illustrated in Figure 1. A total of 372 COVID-19 patients were enrolled after admission from three centers in Guangzhou and Wuhan (Figure 1). All patients with non-severe COVID-19 during hospitalization were followed for more than 15 days after admission. Patients who deteriorated to severe or critical COVID-19 and patients who kept non-severe state were assigned to the severe and non-severe group, respectively. There were no significant differences in age, sex, disease types between the train cohort and validation cohorts (Table 1). COVID-19 group consisted of 161 (85.2%) patients, with a median age of 45 years (range, 33–62 years); 28 patients (14.8%) with a median age of 63.5 years (range, 54.5–72 years) progressed to severe COVID-19. By the end of Feb 25, one patient with severe COVID-19 in the train group died. None of the 189 patients from the train group had a history of exposure to Huanan seafood market in Wuhan, 58 of them (30.7%) had not left Guangzhou recently, but had a close exposure history with COVID-19 patients, and the rest (69.3%) were Wuhan citizens or visited Wuhan recently. Other baseline characteristics in train cohort were shown in Table 2.

**Table 1.** Baseline characteristics of the study cohort

|  | Train cohort<br>(n=189) | Validation cohort 1<br>(n=165) | Validation cohort 2<br>(n=18) | p-value |
| --- | --- | --- | --- | --- |
| Age (years) | 49.0 (35.0, 63.0) | 52.0 (37.0, 64.0) | 41.5 (29.0, 50.0) | 0.05 |
| Gender |  |  |  | 0.77 |
| Female | 101 (53.4%) | 93 (56.4%) | 9 (50.0%) |  |
| Male | 88 (46.6%) | 72 (43.6%) | 9 (50.0%) |  |
| Basic.disease |  |  |  | 0.91 |
| No | 134 (70.9%) | 117 (70.9%) | 14 (77.8%) |  |
| Yes | 55 (29.1%) | 48 (29.1%) | 4 (22.2%) |  |
| Disease type |  |  |  | 0.07 |
| Non-severe | 161 (85.2%) | 125 (75.8%) | 14 (77.8%) |  |
| Severe | 28 (14.8%) | 40 (24.2%) | 4 (22.2%) |  |

“Yes” of Basic.disease means patients with one of the following disease: hypertension, diabetes, cardiovascular disease, chronic respiratory disease, tuberculosis disease.

**Table 2.**
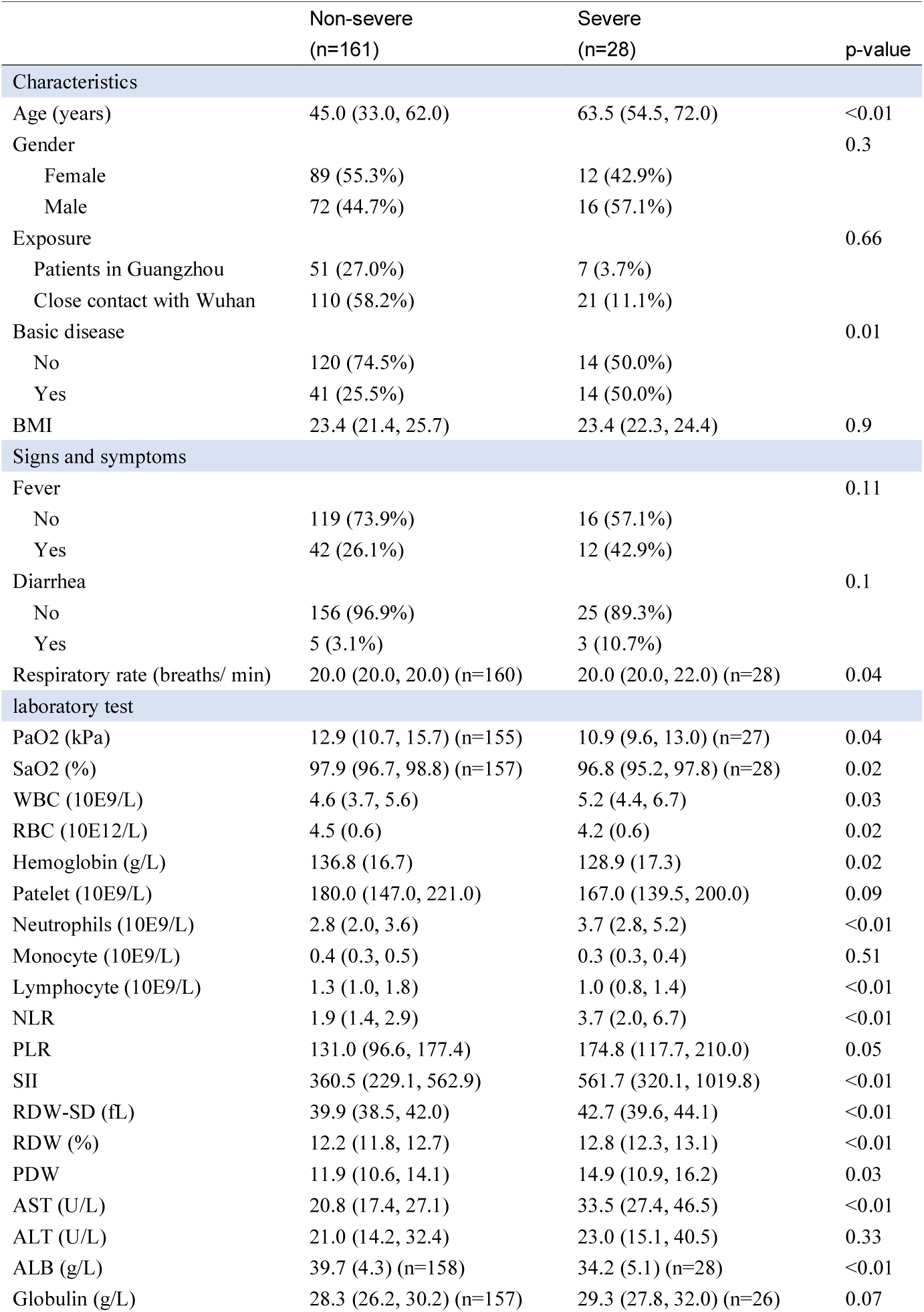
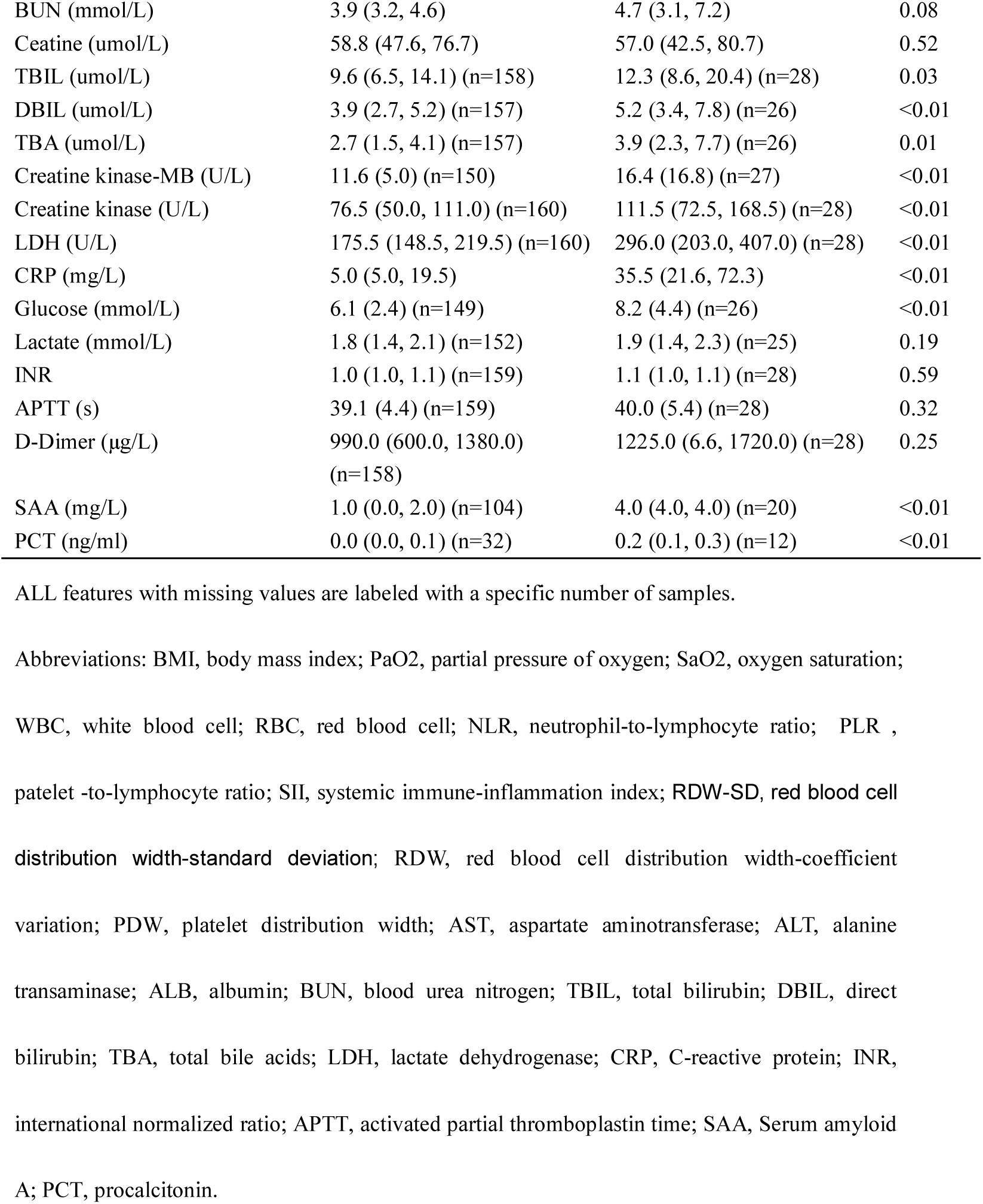
Demographics and characteristics of COVID-19 patients in the train cohort

|  | Non-severe<br>(n=161) | Severe<br>(n=28) | p-value |
| --- | --- | --- | --- |
| **Characteristics** |  |  |  |
| Age (years) | 45.0 (33.0, 62.0) | 63.5 (54.5, 72.0) | <0.01 |
| Gender |  |  | 0.3 |
| Female | 89 (55.3%) | 12 (42.9%) |  |
| Male | 72 (44.7%) | 16 (57.1%) |  |
| Exposure |  |  | 0.66 |
| Patients in Guangzhou | 51 (27.0%) | 7 (3.7%) |  |
| Close contact with Wuhan | 110 (58.2%) | 21 (11.1%) |  |
| Basic disease |  |  | 0.01 |
| No | 120 (74.5%) | 14 (50.0%) |  |
| Yes | 41 (25.5%) | 14 (50.0%) |  |
| BMI | 23.4 (21.4, 25.7) | 23.4 (22.3, 24.4) | 0.9 |
| **Signs and symptoms** |  |  |  |
| Fever |  |  | 0.11 |
| No | 119 (73.9%) | 16 (57.1%) |  |
| Yes | 42 (26.1%) | 12 (42.9%) |  |
| Diarrhea |  |  | 0.1 |
| No | 156 (96.9%) | 25 (89.3%) |  |
| Yes | 5 (3.1%) | 3 (10.7%) |  |
| Respiratory rate (breaths/ min) | 20.0 (20.0, 20.0) (n=160) | 20.0 (20.0, 22.0) (n=28) | 0.04 |
| **laboratory test** |  |  |  |
| PaO2 (kPa) | 12.9 (10.7, 15.7) (n=155) | 10.9 (9.6, 13.0) (n=27) | 0.04 |
| SaO2 (%) | 97.9 (96.7, 98.8) (n=157) | 96.8 (95.2, 97.8) (n=28) | 0.02 |
| WBC (10E9/L) | 4.6 (3.7, 5.6) | 5.2 (4.4, 6.7) | 0.03 |
| RBC (10E12/L) | 4.5 (0.6) | 4.2 (0.6) | 0.02 |
| Hemoglobin (g/L) | 136.8 (16.7) | 128.9 (17.3) | 0.02 |
| Patelet (10E9/L) | 180.0 (147.0, 221.0) | 167.0 (139.5, 200.0) | 0.09 |
| Neutrophils (10E9/L) | 2.8 (2.0, 3.6) | 3.7 (2.8, 5.2) | <0.01 |
| Monocyte (10E9/L) | 0.4 (0.3, 0.5) | 0.3 (0.3, 0.4) | 0.51 |
| Lymphocyte (10E9/L) | 1.3 (1.0, 1.8) | 1.0 (0.8, 1.4) | <0.01 |
| NLR | 1.9 (1.4, 2.9) | 3.7 (2.0, 6.7) | <0.01 |
| PLR | 131.0 (96.6, 177.4) | 174.8 (117.7, 210.0) | 0.05 |
| SII | 360.5 (229.1, 562.9) | 561.7 (320.1, 1019.8) | <0.01 |
| RDW-SD (fL) | 39.9 (38.5, 42.0) | 42.7 (39.6, 44.1) | <0.01 |
| RDW (%) | 12.2 (11.8, 12.7) | 12.8 (12.3, 13.1) | <0.01 |
| PDW | 11.9 (10.6, 14.1) | 14.9 (10.9, 16.2) | 0.03 |
| AST (U/L) | 20.8 (17.4, 27.1) | 33.5 (27.4, 46.5) | <0.01 |
| ALT (U/L) | 21.0 (14.2, 32.4) | 23.0 (15.1, 40.5) | 0.33 |
| ALB (g/L) | 39.7 (4.3) (n=158) | 34.2 (5.1) (n=28) | <0.01 |
| Globulin (g/L) | 28.3 (26.2, 30.2) (n=157) | 29.3 (27.8, 32.0) (n=26) | 0.07 |
| BUN (mmol/L) | 3.9 (3.2, 4.6) | 4.7 (3.1, 7.2) | 0.08 |
| Ceatine (umol/L) | 58.8 (47.6, 76.7) | 57.0 (42.5, 80.7) | 0.52 |
| TBIL (umol/L) | 9.6 (6.5, 14.1) (n=158) | 12.3 (8.6, 20.4) (n=28) | 0.03 |
| DBIL (umol/L) | 3.9 (2.7, 5.2) (n=157) | 5.2 (3.4, 7.8) (n=26) | <0.01 |
| TBA (umol/L) | 2.7 (1.5, 4.1) (n=157) | 3.9 (2.3, 7.7) (n=26) | 0.01 |
| Creatine kinase-MB (U/L) | 11.6 (5.0) (n=150) | 16.4 (16.8) (n=27) | <0.01 |
| Creatine kinase (U/L) | 76.5 (50.0, 111.0) (n=160) | 111.5 (72.5, 168.5) (n=28) | <0.01 |
| LDH (U/L) | 175.5 (148.5, 219.5) (n=160) | 296.0 (203.0, 407.0) (n=28) | <0.01 |
| CRP (mg/L) | 5.0 (5.0, 19.5) | 35.5 (21.6, 72.3) | <0.01 |
| Glucose (mmol/L) | 6.1 (2.4) (n=149) | 8.2 (4.4) (n=26) | <0.01 |
| Lactate (mmol/L) | 1.8 (1.4, 2.1) (n=152) | 1.9 (1.4, 2.3) (n=25) | 0.19 |
| INR | 1.0 (1.0, 1.1) (n=159) | 1.1 (1.0, 1.1) (n=28) | 0.59 |
| APTT (s) | 39.1 (4.4) (n=159) | 40.0 (5.4) (n=28) | 0.32 |
| D-Dimer (µg/L) | 990.0 (600.0, 1380.0) (n=158) | 1225.0 (6.6, 1720.0) (n=28) | 0.25 |
| SAA (mg/L) | 1.0 (0.0, 2.0) (n=104) | 4.0 (4.0, 4.0) (n=20) | <0.01 |
| PCT (ng/ml) | 0.0 (0.0, 0.1) (n=32) | 0.2 (0.1, 0.3) (n=12) | <0.01 |

ALL features with missing values are labeled with a specific number of samples.
Abbreviations: BMI, body mass index; PaO<sub>2</sub>, partial pressure of oxygen; SaO<sub>2</sub>, oxygen saturation;
WBC, white blood cell; RBC, red blood cell; NLR, neutrophil-to-lymphocyte ratio; PLR ,
platelet -to-lymphocyte ratio; SII, systemic immune-inflammation index; RDW-SD, red blood cell
distribution width-standard deviation; RDW, red blood cell distribution width-coefficient
variation; PDW, platelet distribution width; AST, aspartate aminotransferase; ALT, alanine
transaminase; ALB, albumin; BUN, blood urea nitrogen; TBIL, total bilirubin; DBIL, direct
bilirubin; TBA, total bile acids; LDH, lactate dehydrogenase; CRP, C-reactive protein; INR,
international normalized ratio; APTT, activated partial thromboplastin time; SAA, Serum amyloid

**Figure 1.**
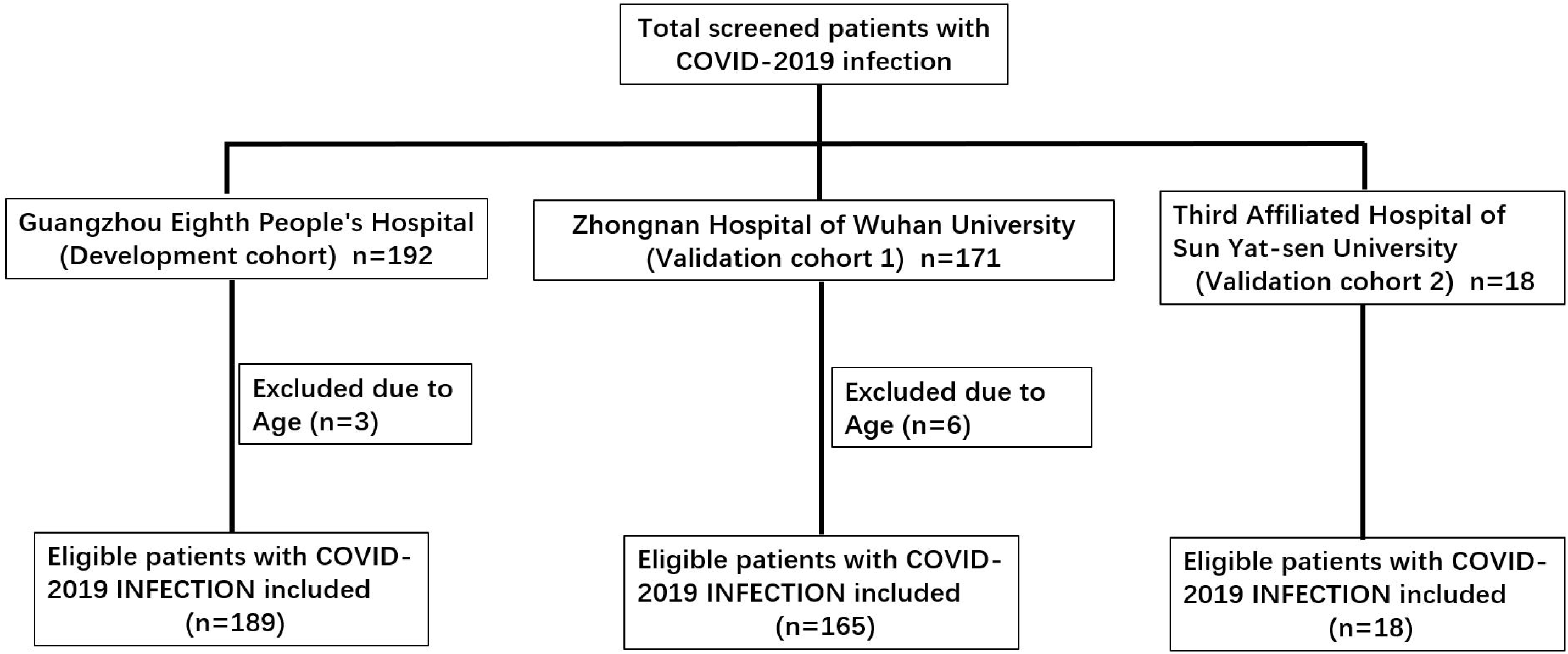
Flow chart of study participants in train and validation groups.

### Prognostic factors of severe COVID-19

A total of 49 features were collected from each patient in the train cohort. After excluding irrelevant and redundant features, 39 features were remained for LASSO regression analysis. The results of the 189 patients showed that age, DBIL, RDW, BUN, CRP, LDH and ALB were predictive factors for severe COVID-19 with maximal AUC (Figure 2A and 2B). Then we built prediction models using logistic regression, decision tree, random forest (RF) and support vector machine (SVM), and evaluated their performance by the receiver operating characteristic curve (ROC) and the precision-recall curve (Supplementary Figure 1). There were no big difference in performance of these models except for decision tree. Therefore, logistic regression model was used for further analysis owing to its high predictive power and interpretability.

**Figure 2.**
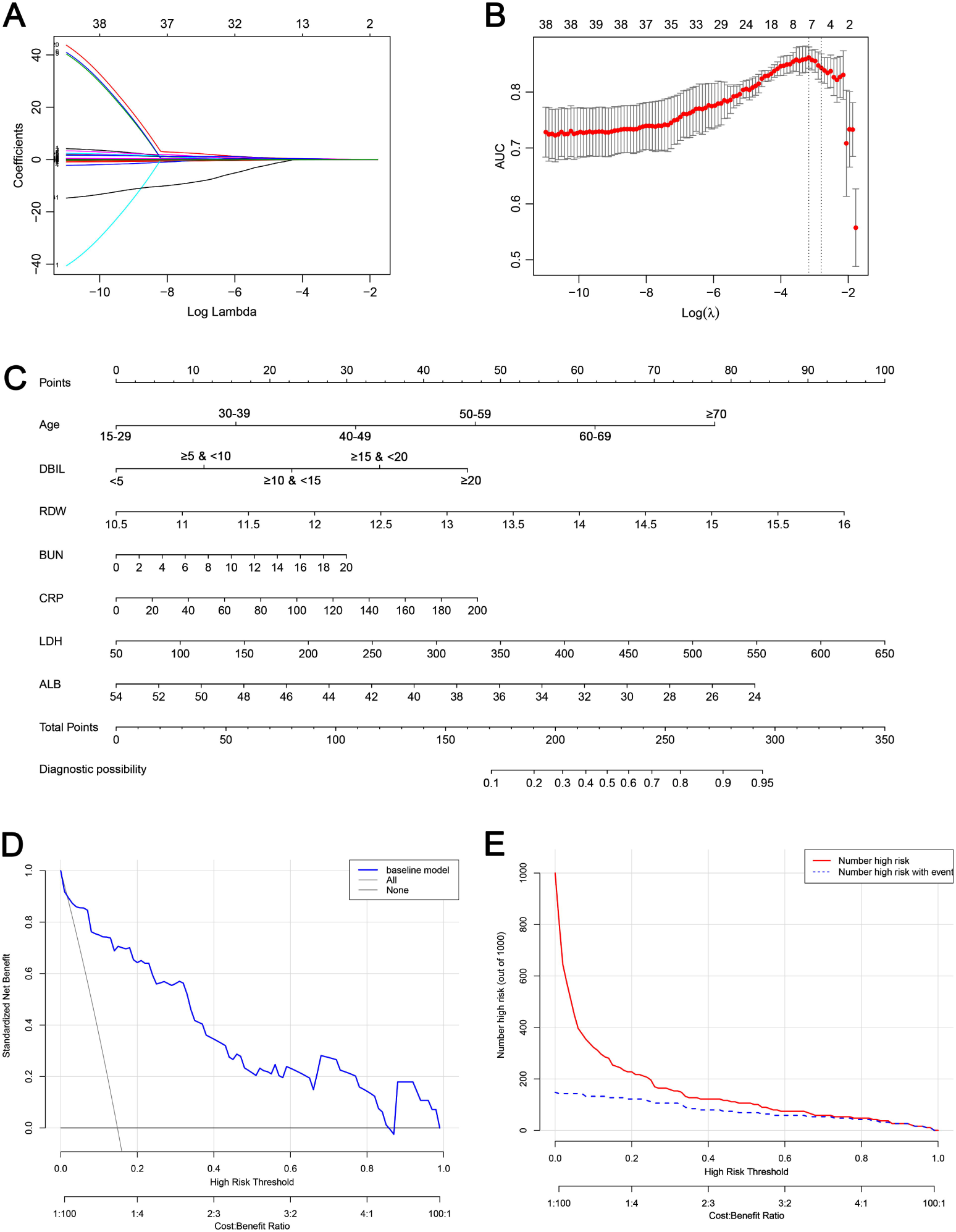
Construction of prediction nomogram in patients with COVID-19. The nomogram composed of age, DBIL, RDW_CV, BUN, CRP, LDH and ALB was developed. (A) LASSO coefficient profiles (y-axis) of the 39 features. The lower x-axis indicated the log (λ). The top x-axis has the average numbers of predictors. (B) Identification of the optimal penalization coefficient (λ) in the LASSO model was peformed via 3-fold cross-validation based on minimum criteria. The area under the receiver operating characteristic (AUC) was plotted verse log (λ). The y-axis indicated AUC. Red dots represent average AUC for each model with a given λ, and vertical bars through the red dots showed the upper and lower values of the AUC. The dotted vertical lines represents the optimal values of λ. When the optimal λ value of 0.042 with log (λ) = - 3.17 was selected, the AUC reached the peak. The upper and lower x-axis indicated the same meaning as in Figure 2A. LASSO, least absolute shrinkage and selection operator. (C) Nomogram predicting the severe COVID-19 probability in patients with COVID-19 infection was plotted. To use this nomogram in clinical management, an individual patient’s value is located on each variable axis, and a line is plotted upward to calculate the number of points received for each variable value. The sum of these scores is located on the Total points axis and draw a line straight down to get the probability of severe COVID-19. (D) Decision curve compares the net clinical benefits of three scenarios in predicting the severe COVID-19 probability: a perfect prediction model (grey line), screen none (horizontal solid black line), and screen based on the nomogram (blue line). (E) Clinical impact curve of the nomogram plot the number of COVID-19 patients classified as high risk, and the number of cases classified high risk with severe NCAP at each high risk threshold. RDW_CV, red blood cell distribution width-coefficient variation; BUN, blood urea nitrogen; DBIL, direct bilirubin; CRP, C-reactive protein; LDH, lactate dehydrogenase; ALB, albumin.

### Nomogram construction

The predictive nomogram that integrated 7 selected features for the incidence of severe COVID-19 in the train cohort is shown (Figure 2C). To evaluate clinical applicability of our risk prediction nomogram, decision curve analysis (DCA) and clinical impact curve analysis (CICA) were performed. The DCA and CICA visually showed that the nomogram had a superior overall net benefit within the wide and practical ranges of threshold probabilities and impacted patient outcomes (Figure 2D and 2E). In Figure 3A and 3B, the calibration plot for severe illness probability showed a good agreement between the prediction by nomogram and actual observation in the train cohort and validation cohort 1, respectively.

**Figure 3.**
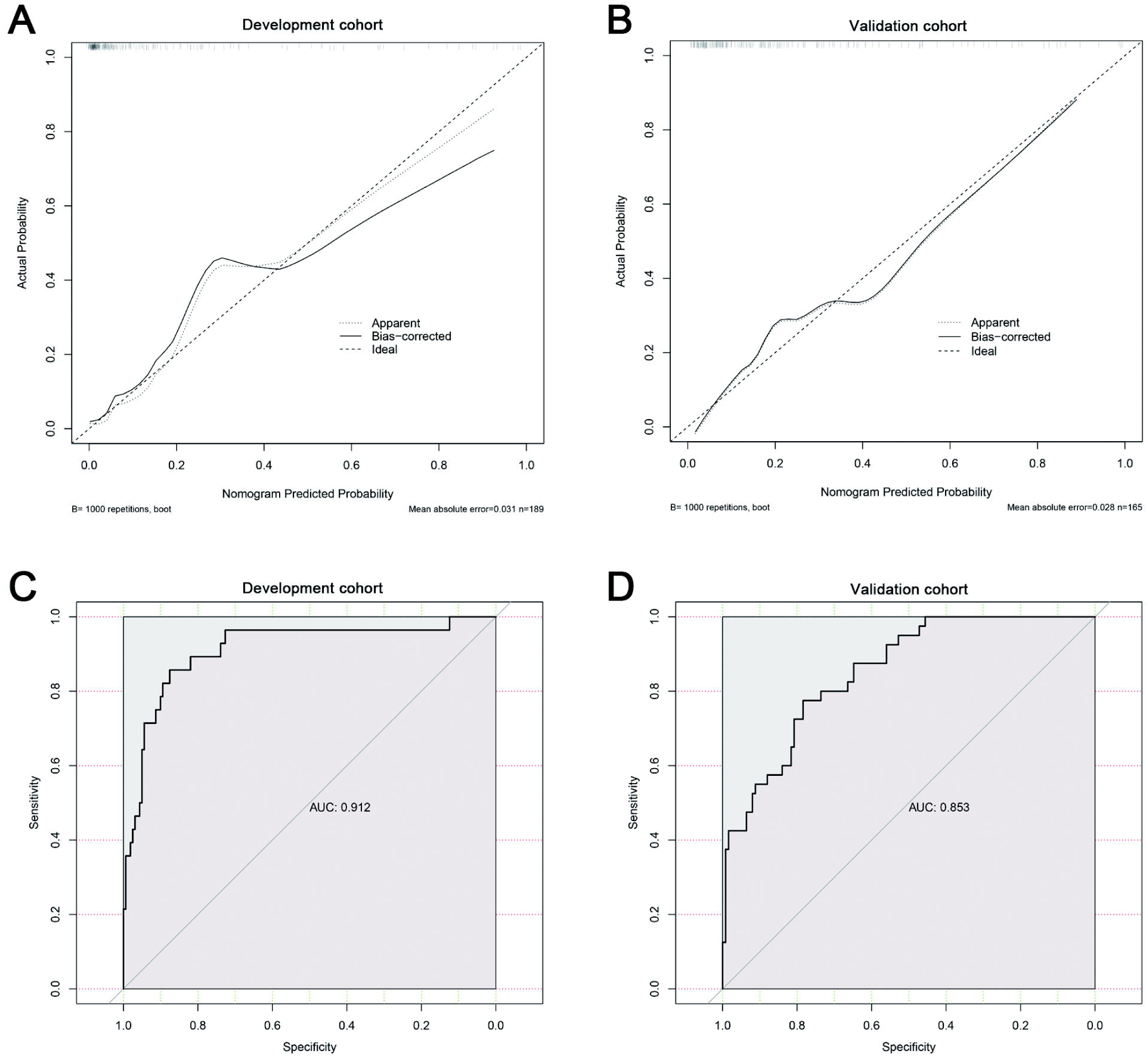
The calibration and ROC curves of the nomogram. The calibration curve and ROC for performance to distinguish individuals with severe COVID-19 from non-severe COVID-19 in the train cohort (A, C) and validation cohort 1 (B, D), respectively.

### Validation of the predictive accuracy of nomogram

In the train cohort, the nomogram had a significantly high AUC 0.912 (95% CI 0.846-0.978) to discriminate individuals with severe COVID-19 from non-severe COVID-19, with a sensitivity of 85.71 % and specificity of 87.58% (Figure 3C, Table 2). Cutpoint R package was used to calculate optimal cutpoints by bootstraping the variability of the optimal cutpoints, which was 188.6358 for our nomogram (corresponding to a threshold probability of 0.190). Then patients in the validation cohorts were divided into the low group (score ≤188.6358) and the high group (score>188.6358) for further analysis. In consistent with the train cohort, in validation cohort 1, AUC was 0.853 for patients with severe COVID-19 versus non-severe COVID-19 with a sensitivity of 77.5 % and specificity of 78.4% (Figure 3D, Table 3). In validation cohort 2, the sensitivity and the specificity of the nomogram were observed to be 75% and 100%, respectively.

**Table 3.** Performance of nomogram for early prediction of severe COVID-19

|  | Severe vs non-severe COVID-19 |  |  |
| --- | --- | --- | --- |
|  | AUC (95% CI) | Sensitivity (%) | Specificity (%) |
| Development cohort<br>(n=189) | 0.912<br>(0.846-0.978) | 85.71 | 87.58 |
| Validation cohort 1<br>(n=165) | 0.853<br>(0.790-0.916) | 77.5 | 78.4 |
| Validation cohort 2<sup>§</sup><br>(n=18) |  | 75 | 100 |

<sup>§</sup> Owing to the limited sample size, AUC was not calculated in validation cohort 2.

## Discussion

Early identification of patients approaching to severe COVID-19 patients will lead to better management and optimal use of medical resources. In this research, we identified older age, higher LDH and CRP, DBIL, RDW, BUN, and lower ALB on admission correlated with higher odds of severe COVID-19. Furthermore, we developed an effective prognostic nomogram composed of 7 features, had significantly high sensitivity and specificity to distinguish individuals with severe COVID-19 from non-severe COVID-19. DCA and CICA further indicated that our nomogram conferred significantly high clinical net benefit, which is of great value for accurate individualized assessment of the incidence of severe COVID-19.

So far, several researches reported some risk factors for severe COVID-19. However, the nomogram could present a quantitive and practical predictor tool for risk stratification of non-severe COVID-19 patients at admission. Though Liu et al developed a nomogram from a single center with a small sample size and no external validation[5], our nomogram has a significantly higher AUC in the train and validation cohorts than Liu’s nomogram (0.912/0.853 vs 0.849). Our nomogram predicted a total of 188.6358 points at a 19.0 % probability threshold, which was close to the prevalence of severe COVID-19 (14.8%) in the training cohort and hence consistent with the reality. This cut-off value may lead to a slight increase of false positive rates but in the setting of this COVID-19 outbreak, a few high false positive rates are acceptable in order to minimize risks of missed diagnosis. Meanwhile, application of the nomogram in the training cohort and validation cohort showed good differentiation with AUC values of 0.912 and 0.853 respectively, as well as high sensitivity and specificity.

Furthermore, only seven easy-access features were included in our nomogram, including older age, higher LDH and CRP, DBIL, RDW, BUN, and lower ALB. Age, NLR and LDH has been reported to be risk factors for severe patients with SARS-CoV-2 infection [3, 5-7]. NLR, a widely used marker for the assessment of system inflammation, was not identified by LASSO as an important feature instead of LDH and CRP, which are associated with the systemic inflammatory response [8]. However, LDH could predict severity of tissue damage in early stage of diseases as an auxiliary marker [9]. These might be reasons why the lasso model did not identified NLR as a more important feature. Consistent with other reports, our results indicate that patients with higher levels of inflammation at admission might be at higher risk for severe COVID-19 as well.

Interestingly, we found RDW was also an important prognostic predictor for severe COVID-19. RDW, one of the numbers or blood cell indices, reflects the variation in the size of RBC (red blood cells), which has been tightly correlated with critical disease [10-12] but negligent in COVID-19. It is a robust predictor of the risk of all cause patient mortality and bloodstream infection in the critically ill [11-14], including acute exacerbation of interstitial pneumonia, ARDS [10, 15]. RDW also can predict prognosis of sepsis, which was tied to poor COVID-19 outcomes-death[16].

The increased RDW in COVID-19 patients may be due to the increased turnover of erythrocytes: 1) Pro-inflammatory states may be responsible for insufficient erythropoiesis with structural and functional alteration of RBC, such as decreased deformability leading to more rapid clearing of RBC. 2) Plasma cytokines such as interleukin 1 (IL-1) and tumor necrosis factor-α (TNF-α) could not only attenuate the renal erythropoietin (EPO) production, but also blunt the erythroid progenitor response to EPO. In addition, INF-γ contributes to apoptosis of the erythroid progenitors and decrease the EPO receptor expression[17]. 3) RBC are dynamic reservoirs of cytokines [18]. Decreased deformability of RBC in severe illness leads to RBC lysis and release of intracellular contents into the circulation[19], including some inflammatory cytokines. This positive feedback could greatly promote the apparent shortened RBC survival and ultimately more morphological variations in cell sizes (i.e., elevated RDW), increased inflammatory response, and lead to severe illness. RDW can be regarded as an index of enhanced patient fragility and higher vulnerability to adverse outcomes [20]. The elevated RDW may explain fatigue experienced by severe COVID-19 patients. Our study has several strengths: first, we provide a practical quantitive prediction tool based on only 7 features which were relatively inexpensive and easy to be obtained directly from the routine blood tests. Second, to guarantee the robustness of the conclusion, we included the data from three centers with a large sample size and validation in independent cohorts. The performance of our nomogram was efficient for clinical practice.

There were some limitations in the study. First, this is a retrospective study, including 372 patients with non-severe COVID-19 on admission. ACE2, the receptor for SARS-COV-2, has been reported to be differentially expressed in different populations[21]. The differences in patient profiles and healthcare might have effect on the performance of nomogram in other populations outside of China. Further studies on different populations with larger patient cohorts are required to verify our findings. Second, some patients are still in hospital and their condition may change with follow-up. The final survival outcome is lacking. Third, the study has not included IgM and IgG antibodies detection. More comprehensive investigations need to be conducted to explain the characteric of the 7 features.

In summary, our data suggest that our nomogram could early identify the severe COVID-19 patients, and RDW was vaulable for prediction of severe diseases. Our nomogram is especially valuable for risk stratification management, which will be helpful for alleviating insufficient medical resources and reducing mortality.

## Data Availability

No additional data available.

## Notes

### Contributors

BH, YLS and FZ designed the study and had full access to all data in the study and take responsibility for the integrity of the data and the accuracy of the data analysis. JG, JYO XPQ, YSJ, YQC and MKT contributed to collect data, analyze data, and write the paper. LXY, JC, MKT and WYX contributed to the statistical analysis. All authors contributed to data interpretation, and reviewed and approved the final version.

## Acknowledgments

This work is funded by the Science and Technology Program of Guangzhou, China (201804010474).

## Conflict of Interest

The author(s) declare(s) that there is no conflict of interest regarding the publication of this paper.

## Notes

### Competing Interest Statement

The authors have declared no competing interest.

